# Implication of backward contact tracing in the presence of overdispersed transmission in COVID-19 outbreak

**DOI:** 10.1101/2020.08.01.20166595

**Authors:** Akira Endo, Centre for the Mathematical Modelling of Infectious Diseases (CMMID) COVID-19 Working Group, Quentin J Leclerc, Gwenan M Knight, Graham F Medley, Katherine E Atkins, Sebastian Funk, Adam J Kucharski

## Abstract

Unlike forward contact tracing, backward contact tracing identifies the source of newly detected cases. This approach is particularly valuable when there is high individual-level variation in the number of secondary transmissions. By using a simple branching process model, we explored the potential of combining backward contact tracing with more conventional forward contact tracing for control of COVID-19.

## Main text

Isolation of symptomatic cases and tracing and quarantine of their contacts is a staple public health control measure, and has the potential to prevent the need for stringent physical distancing policies that result in detrimental impacts on the society (e.g., civil lockdowns) [1,2]. By identifying and quarantining those who have been recently in contact with infected individuals, epidemic control may be achieved without broad restrictions on the general population. Because there is evidence that the number of secondary transmissions of SARS-CoV-2 per case exhibits substantial individual-level heterogeneity (i.e. overdispersion), often resulting in so-called superspreading events [3–5], a large proportion of infections may be linked to a small proportion of original clusters. As a result, finding and targeting originating clusters as well as onwards infection will substantially enhance the effectiveness of tracing methods [6,7]. Here we explore the incremental effectiveness of combining ‘backward’ tracing with conventional ‘forward’ tracing in the presence of overdispersion in SARS-CoV-2 transmission, using a simple branching process model.

## Forward and backward contact tracing

Contact tracing is typically triggered by a confirmed index case identified via symptom-based surveillance. Contacts of this index case are identified via interviews by public health officials (manual contact tracing) or by tracking proximity records on digital devices (digital contact tracing), and asked to quarantine in order to prevent further transmissions. Contact tracing often targets ‘downstream’ individuals, who may have been infected by the index case (‘forward tracing’); i.e. those who have been in contact with the index case after the index case likely became infectious (often assumed as 2 days before illness onset for COVID-19 [8,9]). However, ‘backward tracing’ can also be used to identify the upstream primary case who infected the index case (or a setting or event at which the index case was infected) by retracing history of contact to the likely point of exposure, i.e. up to 14 days prior to symptom onset [10]. If this primary case is identified, a larger fraction of the transmission chain can be detected by forward tracing each of the contacts of this primary case (Figure 1).

**Figure 1.**
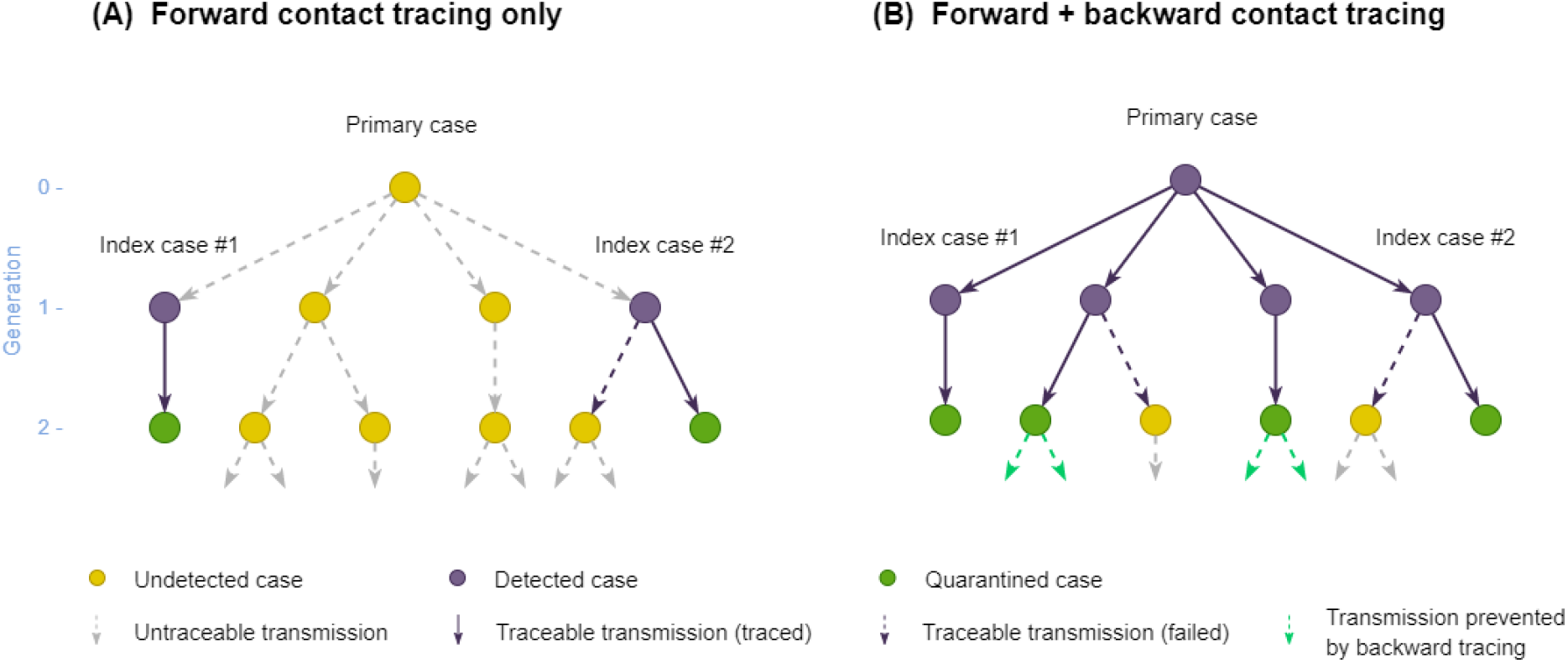
Schematic illustration of forward and backward contact tracing. Two cases (index cases #1 and #2) from a transmission tree originating from an (initially) undetected primary case are assumed to be detected by surveillance. Possible results of contact tracing are shown where (A) only forward tracing is performed; (B) both forward and backward tracing are performed. Some cases may remain undetected because contact tracing can miss cases.

## Overdispersion and the coverage of contact tracing

Unlike forward tracing, backward tracing is more effective when the number of onward transmissions is highly variable, because index cases are disproportionately more likely to have been generated by primary cases who also infected others (an example of the “friendship paradox” [11,12]). We used a branching process model to compare the performance of forward and backward contact tracing triggered by an index case found by symptom-based surveillance. We enumerate generations of transmission chains linked to the index case so that the index case belongs to generation-1 (G1). Backward tracing first identifies the primary case (G0) that infected the index case and then applies forward tracing to those infected by the primary case (G1). We represent the transmission chains of COVID-19 by a branching process where *p*(*x*) denotes the offspring distribution, i.e. the probability mass function of the number of secondary transmissions caused by a single case. If an individual is identified as a primary case, they are more likely to have generated more cases than any random case because the probability that a primary case is identified is proportional to the number of cases it generates. Therefore, the number of offspring of the identified primary case follows 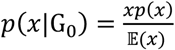, where 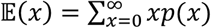. The mean number of G1 cases able to be identified by backward tracing (including the index case) is 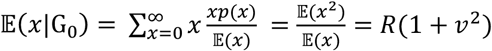, where 𝔼 (*x*) = *R* is the reproduction number and *v* is the coefficient of variation (the standard deviation of *x* divided by its mean). With a high overdispersion (large *v*), backward tracing of the index case can substantially increase the number of G1 cases to trace. Conversely, the mean number of cases that can be identified by forward tracing is *R* regardless of the degree of overdispersion.

When we assume *p*(*x*) follows a negative-binomial distribution [4,13] with an overdispersion parameter *k*, backward tracing on average identifies 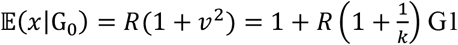 cases. Existing studies suggest *k* for SARS-CoV-2 transmission is small and likely to lie within the range of 0.1-0.5 [4,14,15]. A small *k* indicates that the primary case identified through backward tracing typically generates more secondary cases than does a randomely selected case (𝔼 (*x*) = *R*) (Table 1).

**Table 1.**
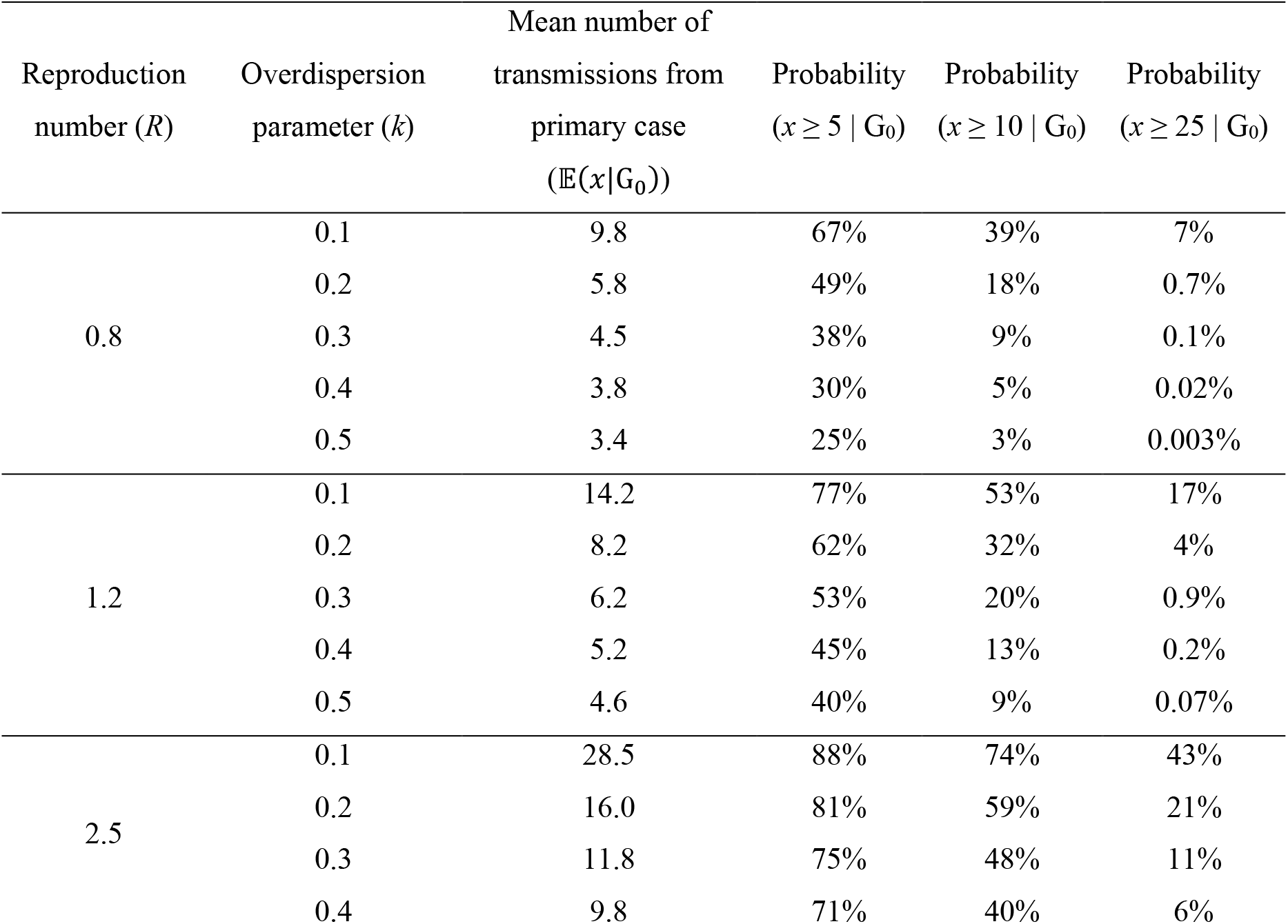
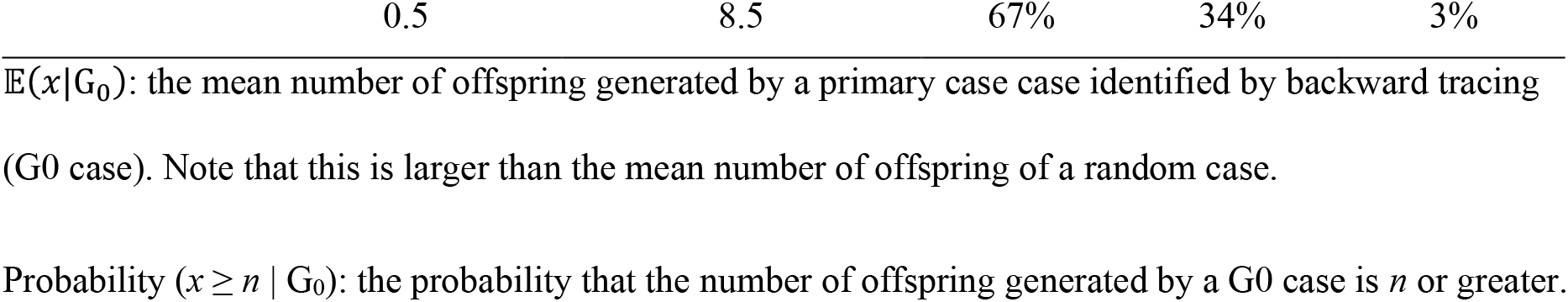
Characteristics of transmissions from a primary case identified by backward contact tracing for different combinations of the reproduction number (*R*) and overdispersion parameter (*k*).

## Simulation of the effectiveness of forward and backward contact tracing

Using our simple branching process model with a negative-binomial offspring distribution, we assessed the potential effectiveness of forward and backward contact tracing. We assumed that contact tracing is triggered by the detection of an index case whose primary case is initially unknown so that our simulation would guide decision making at the operational level (i.e. whether it is worthwhile to implement contact tracing when a case is found). We compared two scenarios: forward tracing only and the combination of forward and backward tracing (Figure 1). In the forward only scenario, G2 cases resulting from an index case are potentially traced and quarantined; in the combined scenario, more G1 cases can be identified through backward tracing of the primary infection and thus a larger number of G2 cases can be traced and quarantined. As the infectious period of G1 cases is likely to have already passed when they are identified by contact tracing because tracing only starts after the index case is confirmed, we assumed that secondary transmissions caused by G1 cases would not be prevented and that only G2 cases successfully traced could be put in quarantine (which confers a relative reduction *c* in infectiousness). To account for potential limitations in the effectiveness of contact tracing, we assumed that the primary case is identified with probability *b* and that each offspring of identified cases are traced with probability *q*. G1 cases not traced may be independently found by symptom-based surveillance; we accounted for such independent case finding with a detection probability *d* (although we excluded backward tracing triggered by these cases from analysis), which is expected to be low due to frequent subclinical infections [16]. We estimated the expected number of G3 cases averted and defined the effectiveness of contact tracing by the relative reduction in the total number of G3 cases. All parameters are listed in Table 2. Detailed methods, the replication code and supplementary figures are reposited on Github (https://github.com/akira-endo/COVID19_backwardtracing).

**Table 2.** Parameter notations and values assumed in simulation

| Parameter | Notation | Assumed value in Figures 2, S1 and S2 |
| --- | --- | --- |
| Reproduction number | $R$ | 1.2, 2.5 |
| Overdispersion parameter | $k$ | 0.2, 0.5 |
| Relative reduction in infectiousness due to quarantine | $c$ | 0.2 – 1.0 |
| Probability of identifying the primary (G0) case by backward tracing | $b$ | 0.5, 0.8 |
| Probability of identifying each offspring of an already identified case | $q$ | 0.0– 1.0 |
| Probability of a G1 case identified by surveillance independently of contact tracing | $d$ | 0.1, 0.2 |

In the forward only scenario, *Rq*(1+*Rd*(1+1/*k*)) G2 cases are traced on average and thus the estimated number of G3 cases averted is *R*^2^*qc*(1+*Rd*(1+1/*k*)). In forward + backward scenario, (1-(1-*d*)(1-*bq*))*R*(1+1/*k*) G1 cases are identified on average in addition to the index case, leading to tracing of *Rq*(1+(1-(1-*d*)(1-*bq*))*R*(1+1/*k*)) G2 cases. *R*^2^*qc*(1+(1-(1-*d*)(1-*bq*))*R*(1+1/*k*)) G3 cases are expected to be averted. Across plausible parameter values, we found that introducing backward tracing in addition to forward tracing increased the effectiveness of contact tracing by a factor of 2-3 (Figures 2, S1 and S2). A higher degree of overdispersion (i.e. small *k*) resulted in a larger absolute number of cases averted by backward tracing (Figures S3 and S4).

**Figure 2.**
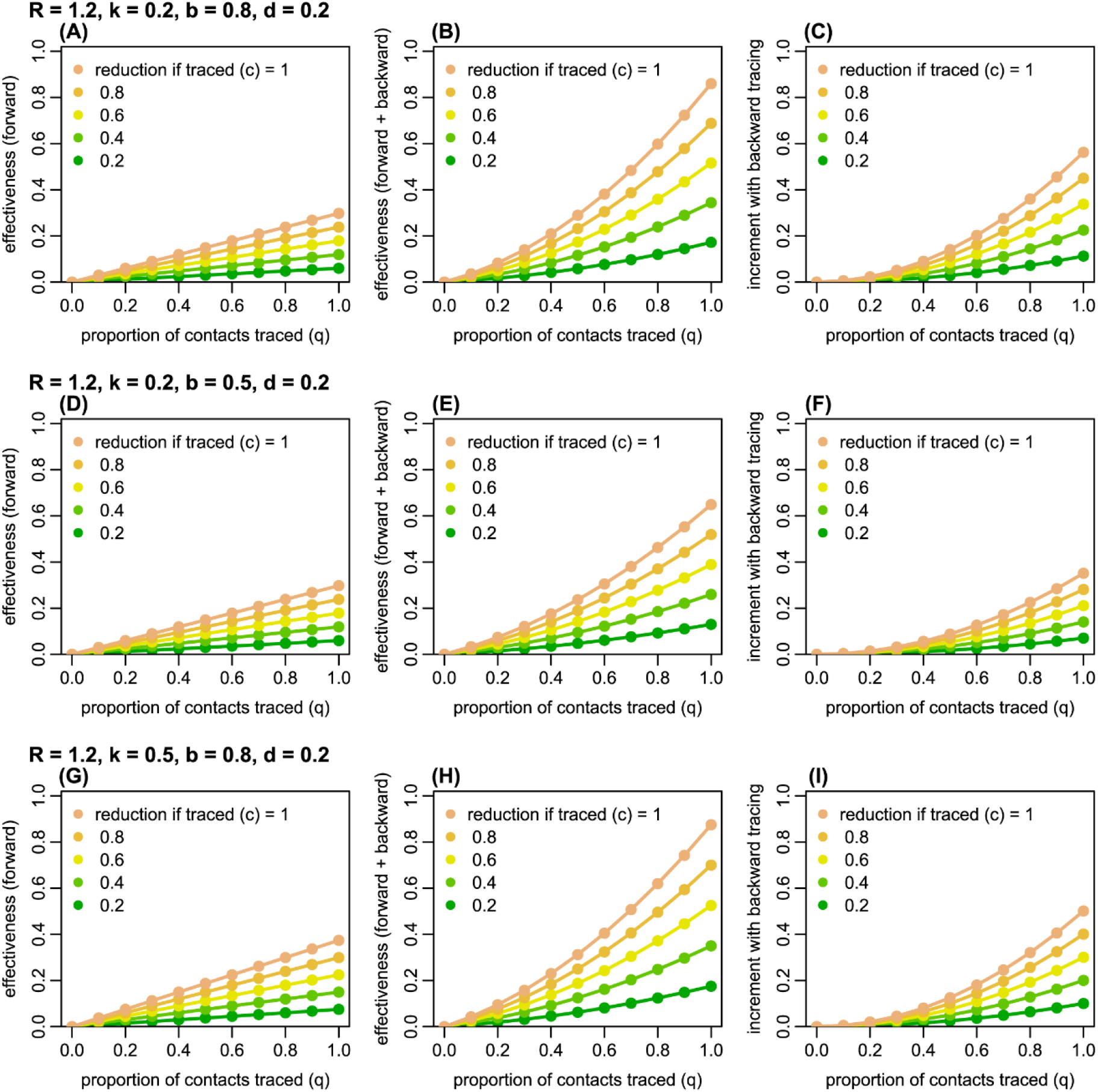
The estimated effectiveness of forward and backward contact tracing for different parameter values. *R*: the reproduction number; *k*: overdispersion parameter; *b*: probability of successful identification of the primary case; *d*: probability of detection of G2 cases independent of contact tracing. Left panels (A, D, G): the effectiveness (the proportion of G3 cases averted) of forward tracing alone; middle panels (B, E, H): the effectiveness of a combination of forward and backward tracing; right panels (C, F, I): incremental effectiveness by combining backward tracing with forward tracing. Colours represent the relative reduction in infectiousness of G2 cases if traced and put in quarantine.

## Discussion

Using a simple branching process model, we showed that backward contact tracing has the potential to identify a large proportion of infections because of the observed overdispersion in COVID-19 transmission. For each index caes detected, forward tracing alone can, on average, identify at most the mean number of secondary infections (i.e. *R*). In contrast, backward tracing increases this maximum number of traceable individuals by a factor of 2-3, as index cases are more likely to come from clusters than a case is to generate a cluster. Furthermore, backward tracing contributes to epidemiological understanding of high-risk settings because transmission events with a common source are more likely to be identified. While standard tracing mostly focuses on forward tracing [8,9], there has been increasing interest in a possible combination of forward and backward tracing to control COVID-19 [7,17]. Our results provide further evidence for this approach by quantifying the possible benefit of backward tracing, especially when the offspring distribution is highly variable, as is the case with SARS-CoV-2.

There are a number of operational challenges to implementing such contact tracing approaches. Since the number of contacts that lead to transmission is likely to be only a fraction of total contacts experienced by detected cases, expanding the coverage of contact tracing may involve a substantial logistical burden [18,19]. With a longer timeline of contact history to be interviewed, recall bias may affect the success rate of backward tracing. In practice, interviewed cases might be asked not only for specific individuals they know to have contacted but also for a history of locations or events visited, as happens during outbreak investigations so that those who were present can be notified and/or tested. Backward tracing can in effect be viewed as an outbreak investigation process in which new cases and their contacts can be routinely linked via their shared exposure events, supported by linkage across epidemiological, diagnostic and quarantine datasets, with additionally identified infections triggering further tracing. Due to the difficulty in determining the direction of transmission, backward tracing may find a cluster of cases linked to an index case rather than a single primary case. However, our results still apply as long as subsequent forward tracing is conducted for the identified cases.

Our model makes some simplifying assumptions. Delays in confirmation and tracing were such that only generation-2 (G2) cases were assumed to be traced and quarantined before becoming infectious. In reality, cases are identified at different points in time and the reduction in infectiousness may be partial if cases are quarantined after becoming infectious (which can be a concern for backward tracing with an additional generation to trace). To allow intuitive comparison, the effectiveness of tracing was measured by the proportion of G3 cases averted given an index case detected by surveillance, and long-term dynamics were not considered.

With these limitations, our results should be considered as a rough estimate suggesting a possible benefit to backward tracing, which should be balanced against finite resources. Because backward tracing is operationally a set of forward tracing measures targeting multiple G1 cases in parallel, additional effectiveness requires a proportional amount of effort, in addition to the ‘overhead’ investigation effort to identify other G1 cases. Cost-effectiveness analysis combined with finer-scale dynamic modelling would help further identify the conditions under which backward tracing is most efficient and feasible.

## Data Availability

No data collection was involved in this study and the replication code is made publicly available.

https://github.com/akira-endo/COVID19_backwardtracing

## Acknowledgement

AE is financially supported by The Nakajima Foundation and The Alan Turing Institute. QJL is supported by Medical Research Council London Intercollegiate Doctoral Training Program studentship (grant no. MR/N013638/1). GMK is supported by UK Medical Research Council (grant: MR/P014658/1). GFM is supported by NTD Modelling Consortium by the Bill and Melinda Gates Foundation (OPP1184344). SF [210758/Z/18/Z] and AJK [206250/Z/17/Z] are sponsored by the Wellcome Trust. The funders had no role in study design, data collection and analysis, decision to publish, or preparation of the manuscript.

## Authors’ contributions

AE, AJK and SF conceptualised the study. AE designed the model and performed the analysis. AE wrote the initial version of the manuscript with inputs from Working Group and QJL, GMK, GFM, KEA, SF and AJK edited it further. All other authors contributed equally and order was assigned randomly.

### CMMID COVID-19 Working Group

Billy J Quilty, Matthew Quaife, Amy Gimma, Charlie Diamond, Rosalind M Eggo, Kiesha Prem, W John Edmunds, Fiona Yueqian Sun, Emily S Nightingale, James W Rudge, Simon R Procter, Rein M G J Houben, Sophie R Meakin, Christopher I Jarvis, James D Munday, Kevin van Zandvoort, Georgia R Gore-Langton, Stéphane Hué, Thibaut Jombart, Damien C Tully, Samuel Clifford, Nicholas G. Davies, Kathleen O’Reilly, Sam Abbott, C Julian Villabona-Arenas, Rachel Lowe, Megan Auzenbergs, David Simons, Nikos I Bosse, Jon C Emery, Yang Liu, Stefan Flasche, Mark Jit, Hamish P Gibbs, Joel Hellewell, Carl A B Pearson, Alicia Rosello, Timothy W Russell, Anna M Foss, Arminder K Deol, Oliver Brady, Petra Klepac

### CMMID COVID-19 Working Group funding statements

Billy J Quilty (NIHR: 16/137/109 & 16/136/46), Matthew Quaife (ERC Starting Grant: 757699, B&MGF: INV-001754), Amy Gimma (Global Challenges Research Fund: ES/P010873/1), Charlie Diamond (NIHR: 16/137/109), Rosalind M Eggo (HDR UK: MR/S003975/1, UK MRC: MC_PC 19065), Kiesha Prem (B&MGF: INV-003174, European Commission: 101003688), W John Edmunds (European Commission: 101003688, UK MRC: MC-PC 19065), Fiona Yueqian Sun (NIHR: 16/137/109), Emily S Nightingale (B&MGF: OPP1183986), James W Rudge (DTRA: HDTRA1-18-1-0051), Simon R Procter (B&MGF: OPP1180644), Rein M G J Houben (ERC Starting Grant: #757699), Sophie R Meakin (Wellcome Trust: 210758/Z/18/Z), Christopher I Jarvis (Global Challenges Research Fund: ES/P010873/1), James D Munday (Wellcome Trust: 210758/Z/18/Z), Kevin van Zandvoort (Elrha R2HC/UK DFID/Wellcome Trust/NIHR, DFID/Wellcome Trust: Epidemic Preparedness Coronavirus research programme 221303/Z/20/Z), Georgia R Gore-Langton (UK MRC: LID DTP MR/N013638/1), Thibaut Jombart (Global Challenges Research Fund: ES/P010873/1, UK Public Health Rapid Support Team, NIHR: Health Protection Research Unit for Modelling Methodology HPRU-2012-10096, UK MRC: MC-PC 19065), Samuel Clifford (Wellcome Trust: 208812/Z/17/Z, UK MRC: MC-PC 19065), Nicholas G. Davies (NIHR: Health Protection Research Unit for Immunisation NIHR200929), Kathleen O’Reilly (B&MGF: OPP1191821), Sam Abbott (Wellcome Trust: 210758/Z/18/Z), Rachel Lowe (Royal Society: Dorothy Hodgkin Fellowship), Megan Auzenbergs (B&MGF: OPP1191821), David Simons (BBSRC LIDP: BB/M009513/1), Nikos I Bosse (Wellcome Trust: 210758/Z/18/Z), Jon C Emery (ERC Starting Grant: #757699), Yang Liu (B&MGF: INV-003174, NIHR: 16/137/109, European Commission: 101003688), Stefan Flasche (Wellcome Trust: 208812/Z/17/Z), Mark Jit (B&MGF: INV-003174; NIHR: 16/137/109, NIHR200929; European Commission: 101003688), Hamish P Gibbs (UK DHSC/UK Aid/NIHR: ITCRZ 03010), Joel Hellewell (Wellcome Trust: 210758/Z/18/Z), Carl A B Pearson (B&MGF: NTD Modelling Consortium OPP1184344, DFID/Wellcome Trust: Epidemic Preparedness Coronavirus research programme 221303/Z/20/Z), Alicia Rosello (NIHR: PR-OD-1017-20002), Timothy W Russell (Wellcome Trust: 206250/Z/17/Z), Oliver Brady (Wellcome Trust: 206471/Z/17/Z), Petra Klepac (Royal Society: RP\EA\180004, European Commission: 101003688)

## References

1. Ferretti L, Wymant C, Kendall M, Zhao L, Nurtay A, Abeler-Dörner L, et al. Quantifying SARS-CoV-2 transmission suggests epidemic control with digital contact tracing. Science (80-). 2020;368:eabb6936. doi:10.1126/science.abb6936

2. Kucharski AJ, Klepac P, Conlan AJK, Kissler SM, Tang ML, Fry H, et al. Effectiveness of isolation, testing, contact tracing, and physical distancing on reducing transmission of SARS-CoV-2 in different settings: a mathematical modelling study. Lancet Infect Dis. 2020. doi:10.1016/S1473-3099(20)30457-6

3. Liu Y, Eggo RM, Kucharski AJ. Secondary attack rate and superspreading events for SARS-CoV-2. Lancet. 2020. doi:10.1016/S0140-6736(20)30462-1

4. Endo A, Abbott S, Kucharski AJ, Funk S. Estimating the overdispersion in COVID-19 transmission using outbreak sizes outside China. Wellcome Open Res. 2020;5:67. doi:10.12688/wellcomeopenres.15842.3

5. Leclerc QJ, Fuller NM, Knight LE, Funk S, Knight GM. What settings have been linked to SARS-CoV-2 transmission clusters? Wellcome Open Res. 2020;5:83. doi:10.12688/wellcomeopenres.15889.2

6. Klinkenberg D, Fraser C, Heesterbeek H. The Effectiveness of Contact Tracing in Emerging Epidemics. Getz W, editor. PLoS One. 2006;1:e12. doi:10.1371/journal.pone.0000012

7. Bradshaw WJ, Alley EC, Huggins JH, Lloyd AL, Esvelt KM. Bidirectional contact tracing is required for reliable COVID-19 control. medRxiv. 2020; 2020.05.06.20093369. doi:10.1101/2020.05.06.20093369

8. World Health Organization. Contact tracing in the context of COVID-19: Interim guidance. 2020. Available: https://www.who.int/publications/i/item/contact-tracing-in-the-context-of-covid-19

9. Centers for Disease Control and Prevention. Health Departments: Interim Guidance on Developing a COVID-19 Case Investigation & Contact Tracing Plan. 2020. Available: https://www.cdc.gov/coronavirus/2019-ncov/downloads/case-investigation-contact-tracing.pdf

10. Backer JA, Klinkenberg D, Wallinga J. Incubation period of 2019 novel coronavirus (2019-nCoV) infections among travellers from Wuhan, China, 20–28 January 2020. Eurosurveillance. 2020;25. doi:10.2807/1560-7917.ES.2020.25.5.2000062

11. Salathe M, Kazandjieva M, Lee JW, Levis P, Feldman MW, Jones JH. A high-resolution human contact network for infectious disease transmission. Proc Natl Acad Sci .2010;107:22020–22025. doi:10.1073/pnas.1009094108

12. Allard A, Moore C, Scarpino S V., Althouse BM, Hébert-Dufresne L. The role of directionality, heterogeneity and correlations in epidemic risk and spread. 2020. Available: http://arxiv.org/abs/2005.11283

13. Lloyd-Smith JO, Schreiber SJ, Kopp PE, Getz WM. Superspreading and the effect of individual variation on disease emergence. Nature. 2005;438:355–359. doi:10.1038/nature04153

14. Lau MSY, Grenfell B, Nelson K, Lopman B. Characterizing super-spreading events and age-specific infectivity of COVID-19 transmission in Georgia, USA. medRxiv. 2020; 2020.06.20.20130476. doi:10.1101/2020.06.20.20130476

15. Adam D, Wu P, Wong J, Lau E, Tsang T, Cauchemez S, et al. Clustering and superspreading potential of severe acute respiratory syndrome coronavirus 2 (SARS-CoV-2) infections in Hong Kong. Prepr (Version 1) available Res Sq. 2020. doi:https://doi.org/10.21203/rs.3.rs-29548/v1

16. Oran DP, Topol EJ. Prevalence of Asymptomatic SARS-CoV-2 Infection. Ann Intern Med. 2020; M20–3012. doi:10.7326/M20-3012

17. Scientific Pandemic Influenza Group on Modelling Operational sub-group. SPI-M-O: Consensus Statement on COVID-19, 3 June 2020. 2020. Available: https://assets.publishing.service.gov.uk/government/uploads/system/uploads/attachment_data/file/897526/S0471_SAGE_40_200603_SPI-M-O_Consensus_Statement.pdf

18. Keeling MJ, Hollingsworth TD, Read JM. Efficacy of contact tracing for the containment of the 2019 novel coronavirus (COVID-19). J Epidemiol Community Health .2020; jech-2020-214051. doi:10.1136/jech-2020-214051

19. Hellewell J, Abbott S, Gimma A, Bosse NI, Jarvis CI, Russell TW, et al. Feasibility of controlling COVID-19 outbreaks by isolation of cases and contacts. Lancet Glob Heal. 2020;8:e488–e496. doi:10.1016/S2214-109X(20)30074-7

